# Cardiovascular risk in aging adults with double deficits in social support: a gender-sensitive, cross-sectional analysis of the CLSA cohort

**DOI:** 10.1101/2024.08.18.24312193

**Authors:** Annalijn I. Conklin, Abdollah Safari, Gerry Veenstra, Nadia A. Khan

## Abstract

**Background:** Synergistic effects of different forms of social support (informational, tangible, emotional and belonging) on cardiovascular disease risk factors (CVRF) in aging women versus men is unknown.

**Aim:** To quantify gender differences in the additive and multiplicative associations of four different forms of social support (informational, tangible, emotional and belonging) with cardiovascular disease risk factors (CVRF) in aging adults.

**Methods:** Baseline data from 28,779 adults (45-85 years) in the Canadian Longitudinal Study on Aging Comprehensive cohort (2012-15) with four self-reported social support measures and measured body mass index, waist circumference and blood pressure. We used stratified multivariable principal component regression (PCR) and then post-estimated calculation of adjusted means and 95% CIs.

**Results:** Among women, reporting low availability of two types of social support was consistently associated with the highest adjusted mean levels of WC and BMI. However, lack of two types of support was less clearly or consistently associated with anthropometric measures among men. The highest adjusted mean SBP levels among women were seen for women with low availability of informational support. Among men, low-low combinations of two social supports did not consistently correspond to the highest adjusted mean SBP levels. Results for DBP were null and showed no gender differences. Sensitivity analyses revealed two significant interaction effects of tangible and belonging supports for SBP and WC among women.

**Conclusion:** Findings showed women with two deficits in social supports had the worse CVRF profiles than one social support deficit. Women’s worse CVRF profiles were seen for deficit combinations that included low informational support. We did not, however, find antagonistic/synergistic effects of social support on CVRFs. Heart health care and prevention for aging women would benefit from ensuring informational support with other supports is available.

## Introduction

Elevated blood pressure (BP) is a vital sign of health and a major cardiovascular risk factor (CVRF) that is more prevalent among women than men in later life.^1^ Blood pressure rises more with age among women than among men which may be due to sex/gender and age differences in the changing balance of vasodilating and vasoconstricting adrenergic receptor tone.^2^ Both hypertension and obesity prevalence contribute to the global burden of disease and are strong physiological determinants that contribute to unhealthy ageing.^3^ The annual costs of hypertension alone were expected to be $20.5 billion in 2020 in Canada.^4^ The burden of obesity has more than doubled in Canada since the 1970s, with faster rates observed among women in extreme classes of obesity.^5^ Preventing and treating CVRFs is a public health and policy priority in Canada and elsewhere.^6^

Social isolation during the early restrictive stages of the COVID-19 pandemic was shown to increase BP among emergency admissions in Argentina.^7^ Social support is known to have beneficial or adverse effects on cardiac and metabolic health^8^ and is directly associated with psychological wellbeing.^9,10^ Higher perceived social support has increased quality of life in cardiovascular patients in South Asia, especially in those with high self-efficacy.^11^ Although high social support buffers against life stress (stress buffering theory), research also shows that high social support can combine with high social undermining (i.e. efforts by group members to intentionally hinder another member’s success) to negatively impact health.^12^ Furthermore, there may be gender differences in the health buffering role of supportive networks;^13^ for example, synergistic interactions of social support and job control on depression and insomnia appear stronger among employed women than among men.^14^ Recent research indicates that greater informational and emotional support are separately linked to lower blood pressure, more strongly so for women than for men;^15^ and, similarly, different social supports are associated with obesity in gender-specific kinds of ways.^16^

As a multidimensional construct, social support has traditionally been measured using a summary index (e.g. MOS score),^17^ and so the interplay of different social supports is seldom considered. Whether the absence of one type of social support can be mitigated by the presence of another type of support or whether double deficits in supports have synergistic effects for women or men is unknown. This prospective study therefore aimed to investigate effects of multiple social supports and their interactions in relation to major CVRFs in an aging population using a gender-based analysis. We hypothesized that dyadic interactions between the four types of social support (informational, tangible, emotional and belonging) would be synergistically associated with the CVRFs wherein low levels of two forms of support correspond to inordinately worse measures of CVRFs. We also hypothesized that these synergistic effects would be more pronounced among women than among men.

## Methods

### Study design and population

We used baseline data (2011-15) of the population-based Canadian Longitudinal Study on Aging (CLSA) Comprehensive cohort that provided information on physical, social and psychological factors as well as objective measures of CVRFs from approximately 30,000 respondents aged 45-85 years living in the community.^18^ Excluded from CLSA Comprehensive cohort were people not speaking English or French; having cognitive impairment; with temporary visa or transitional health coverage; living in the three territories, some remote regions, on federal First Nations reserves and other First Nations settlements in the provinces, Canadian Armed Forces full-time members, and living in institutions; and participants not completing both the in-home interview and data collection site visit at baseline.^19^ Our available sample included participants with complete data on perceived availability of social support, covariables, anthropometry and blood pressure (n=28,779). All participants gave written informed consent and the study was approved by the *[blinded for review]* Behavioural Research Ethics Board (H19-00971). CLSA data were fully anonymized for research purposes and were accessed on 31-07-2019.

### CVD risk factors (CVRFs)

We used two anthropometric measures (body mass index (kg/m^2^) and waist circumference (cm)) and two cardiovascular measures (systolic blood pressure (SBP) and diastolic blood pressure (DBP)) as important indicators of cardiovascular disease (CVD) risk, and established physiological determinants of ageing.^20^ We used clinically measured height (m) and weight (Kg) to calculate body mass index (BMI, Kg/m^2^). We used the last five of six measurements to calculate a mean value for SBP and DBP for each participant; clinical measures were taken with the BpTRU™ BPM200 Blood Pressure Monitor.^19^ We also used clinically relevant secondary outcomes of general obesity (BMI ≥ 30 kg/m^2^), central obesity (WC ≥ 88 cm for women and WC ≥ 102 cm for men) and hypertension (≥ 140/90 mmHg (≥ 130/80 for those with diabetes), self-reported diagnosis or use of hypertension medication).

### Four types of social support

Participants self-reported the perceived availability of emotional, information, tangible, and belonging support based on questions taken from the Medical Outcomes Study (MOS) Social Support Survey,^17^ that provide five response options (none, a little, some, most and all of the time).^19^ Questions assessing *emotional* social support asked about having someone: to listen when needing to talk, to confide in about oneself or problems, to share their most private worries and fears with, to love and make them feel wanted and who shows love and affection, and who hugs them. Questions on *informational* support included having someone to give advice about a crisis, to give information in order to help, to turn to for suggestions about how to deal with a personal problem, and someone whose advice is really wanted. Questions about *tangible* social support concerned availability of help if confined to a bed, someone to take to the doctor, to prepare meals if unable to do so alone, or to help with daily chores if sick. And questions regarding *belonging* support reflected having someone to have a good time with, to relax with, to do things to help get one’s mind off things, to do something enjoyable with, and who understands one’s problems.

### Covariables

We included a parsimonious set of covariables: self-reported age and age squared, smoking status (ever/never), province and education (less than secondary school; secondary school; some post-secondary education including degree/diploma; university degree). Our selection of covariables was informed by existing literature on factors known to correlate with our exposure (social ties) and our outcome^21,22^ and were not mediators on the causal pathway (e.g. health behaviours). Nevertheless, results were checked for robustness to other factors that might be potential confounders: chronic conditions (hyper-and hypothyroidism; rheumatoid arthritis; asthma; CVD; cancer, osteoporosis; diabetes; Parkinson’s; and stroke; or relevant medications); marital status; lifestyle factors (e.g., weekly alcohol intake; daily servings of fruits and vegetables; sleep duration and quality; amount of daily physical activity); psychological factors (Center for Epidemiological Studies Depression scale (CESD), life satisfaction and depression medication), blood pressure (systolic and diastolic, Hg mm) or BMI (depending on the outcome modelled). Sex-specific risk factors related to women’s reproductive status (i.e., number of biological children (numeric), menopause status (Y/N) and hormone replacement therapy use (ever/never)) were also tested in sensitivity analyses.

### Statistical analysis

Descriptive statistics (mean (SD) or proportions) were used to summarize socio-demographic characteristics and crude levels of cardio-metabolic outcomes in relation to the four functional social ties. As the CLSA Comprehensive cohort survey weights represented only Canadian population residing within 25 km of CLSA’s Data Collection Site they were not used here. Different statistical tests were used to measure the strength of relationships between the functional tie variables.

A sequence of multivariate linear and logistic regression models was used to capture additive and multiplicative associations of the different functional ties with adiposity and cardiovascular outcomes by gender. Stratified multivariable regression models were used to report estimates separately for (cisgender) women and men. The models for each outcome simultaneously accounted for known confounders (see covariables), functional ties, and two-way interactions between multiple functional ties. Due to the high multicollinearity between the functional ties and also their interactions (variance inflation factors > 5), we used principal component regression (PCR) models to analyze their interactive associations.^23^ Three main steps are involved in fitting a PCR model: 1) Compute principal components (PCs) on correlated covariates (functional ties and their interactions); 2) Run a regression model on PCs and other independent cofounders; and 3) Back transform the coefficients’ estimate of the PCs in the regression model to the coefficient of original correlated predictors by using the PCs’ scores. A subset of PCs will be used in the regression model in Step 2 in order to avoid multicollinearity issues between the correlated covariates. We employed R-squared and root mean squared error of prediction (RMSEP) criteria to decide how many PCs should be dropped from the regression model in Step 2.^24^ In most cases, only the last one or two PCs were removed from the regression models.

Adjusted mean levels and 95% confidence intervals (CI95) of continuous outcomes (WC, BMI, SBP and DBP) were calculated based on post-estimation analysis for women and men and displayed visually for ease of interpretation. Adjusted means of different combinations of levels of each functional tie pair were also compared using Tukey adjusted p-values for multiple testing. Sensitivity analyses were used to test the robustness of the interactive associations to additional confounding. Analyses were carried out in R (a language and environment for statistical computing. R Foundation for Statistical Computing, Vienna, Austria, 2020).

## Results

The mean age of participants was 63 (SD 10) years with 50.8% women (Table 1). Educational differences were seen between those with high versus low social support of each type. A higher proportion of Canadian adults with low social support were generally obese, centrally obese or had hypertension. Aging adults with low social support had higher mean BMI and mean SBP than adults with high social support.

**Table 1.** Descriptive characteristics across four types of social support in the CLSA cohort at baseline (2012-15).

|  | N | Women | Mean<br>(SD)<br>age | Highest<br>education | Non-<br>smoker | Mean<br>(SD)<br>WC | Central<br>obesity | Mean<br>(SD)<br>BMI | General<br>obesity | Mean<br>(SD)<br>SBP | Mean<br>(SD)<br>DBP | Hyper-<br>tension |
| --- | --- | --- | --- | --- | --- | --- | --- | --- | --- | --- | --- | --- |
| **Total** | 28,779 | 14627<br>(51%) | 62.73<br>(10.18) | 13123<br>(46%) | 9107<br>(32%) | 94.12<br>(14.59) | 12630<br>(44%) | 28.03<br>(5.37) | 8404<br>(29%) | 120.98<br>(16.63) | 73.87<br>(9.98) | 14171<br>(49%) |
| **Informational support** |  |  |  |  |  |  |  |  |  |  |  |  |
| High (20) | 7650 | 4024<br>(53%) | 61.58<br>(9.87) | 4014<br>(52%) | 2681<br>(35%) | 93.13<br>(14.38) | 3143<br>(41%) | 27.78<br>(5.20) | 2084<br>(27%) | 120.20<br>(16.31) | 73.99<br>(9.68) | 3454<br>(45%) |
| Low (4-19) | 21,129 | 10603<br>(50%) | 63.15<br>(10.26) | 9109<br>(43%) | 6426<br>(30%) | 94.48<br>(14.65) | 9487<br>(45%) | 28.12<br>(5.42) | 6320<br>(30%) | 121.26<br>(16.74) | 73.83<br>(10.09) | 10717<br>(51%) |
| **Tangible support** |  |  |  |  |  |  |  |  |  |  |  |  |
| High (20) | 8685 | 3827<br>(44%) | 62.47<br>(9.81) | 4517<br>(52%) | 2884<br>(33%) | 94.52<br>(14.29) | 3636<br>(42%) | 27.92<br>(5.11) | 2391<br>(28%) | 120.68<br>(16.18) | 73.97<br>(9.71) | 4190<br>(48%) |
| Low (4-19) | 20,094 | 10800<br>(54%) | 62.85<br>(10.34) | 8606<br>(43%) | 6223<br>(31%) | 93.95<br>(14.72) | 8994<br>(45%) | 28.07<br>(5.47) | 6013<br>(30%) | 121.11<br>(16.82) | 73.83<br>(10.10) | 9981<br>(50%) |
| **Emotional support** |  |  |  |  |  |  |  |  |  |  |  |  |
| High (35) | 7125 | 3552<br>(50%) | 61.57<br>(9.88) | 3628<br>(51%) | 2442<br>(34%) | 93.61<br>(14.45) | 2943<br>(41%) | 27.85<br>(5.18) | 1980<br>(28%) | 120.25<br>(16.23) | 74.02<br>(9.74) | 3238<br>(45%) |
| Low (7-34) | 21,654 | 11075<br>(51%) | 63.12<br>(10.25) | 9495<br>(44%) | 6665<br>(31%) | 94.29<br>(14.63) | 9687<br>(45%) | 28.08<br>(5.42) | 6424<br>(30%) | 121.22<br>(16.75) | 73.83<br>(10.06) | 10933<br>(50%) |
| **Belonging support** |  |  |  |  |  |  |  |  |  |  |  |  |
| High (20) | 8026 | 3913<br>(49%) | 61.92<br>(9.82) | 4003<br>(50%) | 2643<br>(33%) | 93.90<br>(14.38) | 3365<br>(42%) | 27.93<br>(5.16) | 2255<br>(28%) | 120.65<br>(16.21) | 74.06<br>(9.68) | 3744<br>(47%) |
| Low (4-19) | 20,753 | 10714<br>(52%) | 63.05<br>(10.30) | 9120<br>(44%) | 6464<br>(31%) | 94.20<br>(14.67) | 9265<br>(45%) | 28.06<br>(5.44) | 6149<br>(30%) | 121.11<br>(16.79) | 73.80<br>(10.09) | 10427<br>(50%) |

DBP, diastolic blood pressure; SBP, systolic blood pressure. Hypertension was determined based on SBP ≥ 140 mm Hg and/or DBP ≥ 90 mm Hg (except patients with diabetes, ≥ 130/80), self-reported diagnosis of hypertension, or use of medication for hypertension. Highest education level: minimum having a Bachelor's degree from a university, or higher.

### Additive and interactive associations of dual social supports with anthropometry, by gender

Figure 1 shows that low availability of two types of support was associated with the highest levels of BMI among women. Women with low informational support and low tangible support showed the greatest difference in adjusted mean BMI (27.95 kg/m^2^ [27.93, 27.97]) compared to women with high informational support and high tangible support (27.34 kg/m^2^ [27.30, 27.38]) (data in Supplementary Table S1). All low-low combinations of support were linked to the highest adjusted mean BMI levels among women (range: 27.92 to 27.95 kg/m^2^). There was a linear trend observed for the combination of informational support with each of the three other types of support, suggesting that the absence of either informational support or another support had similar adverse effects on BMI among women. The pattern of an additive effect of lack of two supports on BMI was less clear among men. Specific combinations of high belonging support or high emotional support with low availability of a second type of support were associated with the highest adjusted mean BMI levels in the range of 28.45 to 28.64 kg/m^2^; that is, most low-low, or even high-high, combinations of social supports were not associated with, respectively, the highest or lowest adjusted mean BMI levels among men (Figure 1 and Supplementary Table S1).

**Figure 1.**
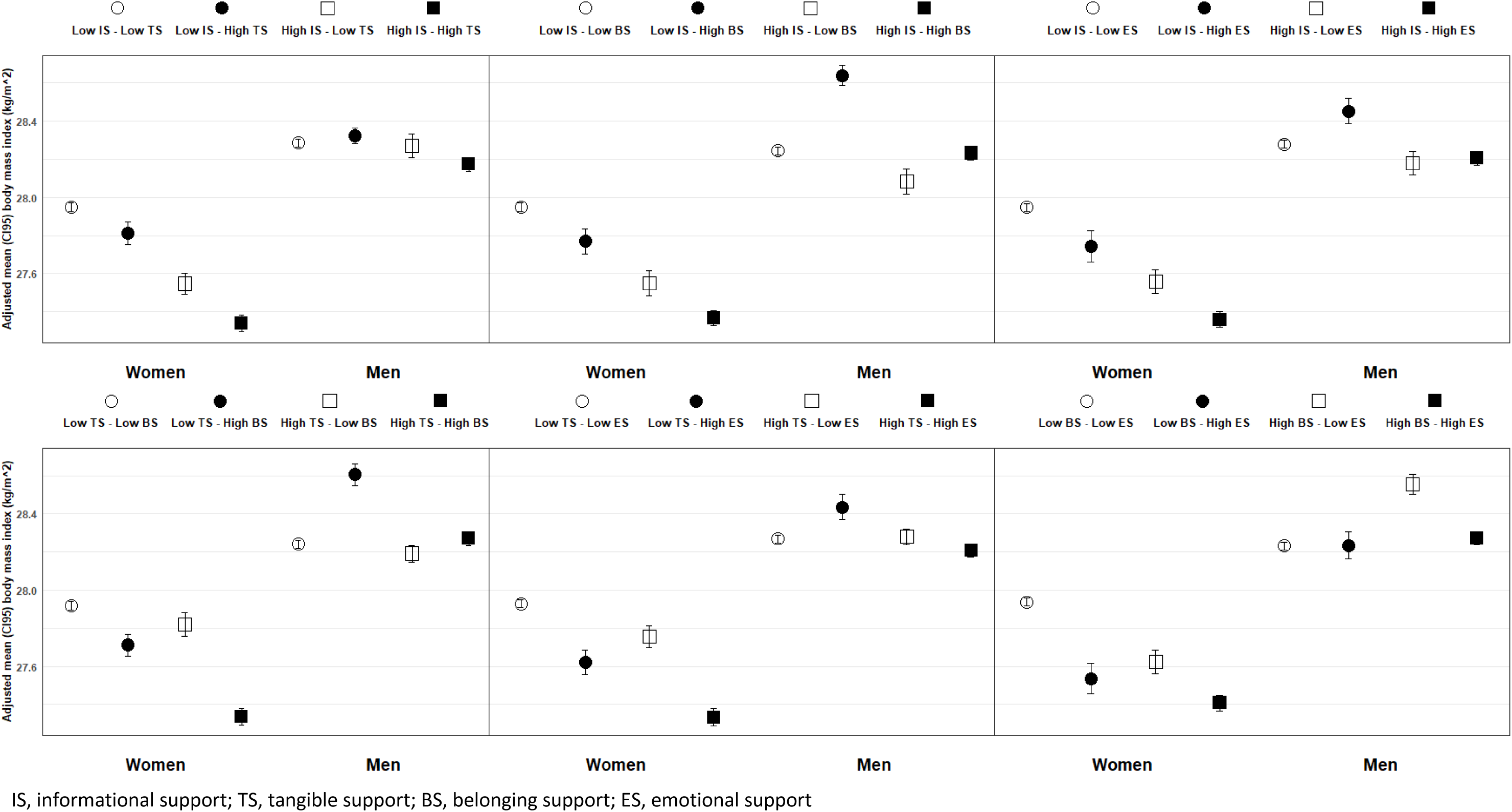
Adjusted mean BMI for the interactive associations of four types of perceived social support in women and men

Similar gender-specific, but much weaker, associations between two social supports were seen for adjusted mean levels of WC among women and men (Figure 2 and Supplementary Table S1). Again, all low-low combinations were associated with the highest adjusted mean WC levels among women (range: 88.62 to 88.71 cm), whereas all high-high combinations of social supports were associated with the lowest adjusted mean WC levels among women (range: 86.96 to 87.06 cm). The greatest difference in adjusted mean WC was seen among women with low informational support and low emotional support (88.69 cm [88.62, 88.76]) compared to women with high informational and high emotional supports (86.88 cm [86.75, 87.01]). Similar to BMI, a lack of either informational support or another support had similarly adverse effects on WC among women. By contrast, men showed a mostly flat pattern of association between different low-high combinations of two social supports and mean WC with average WC levels for all high-high combinations ranging from 99.73 cm to 99.99 cm and all low-low combinations ranging from 100.02 cm to 100.32 cm.

**Figure 2.**
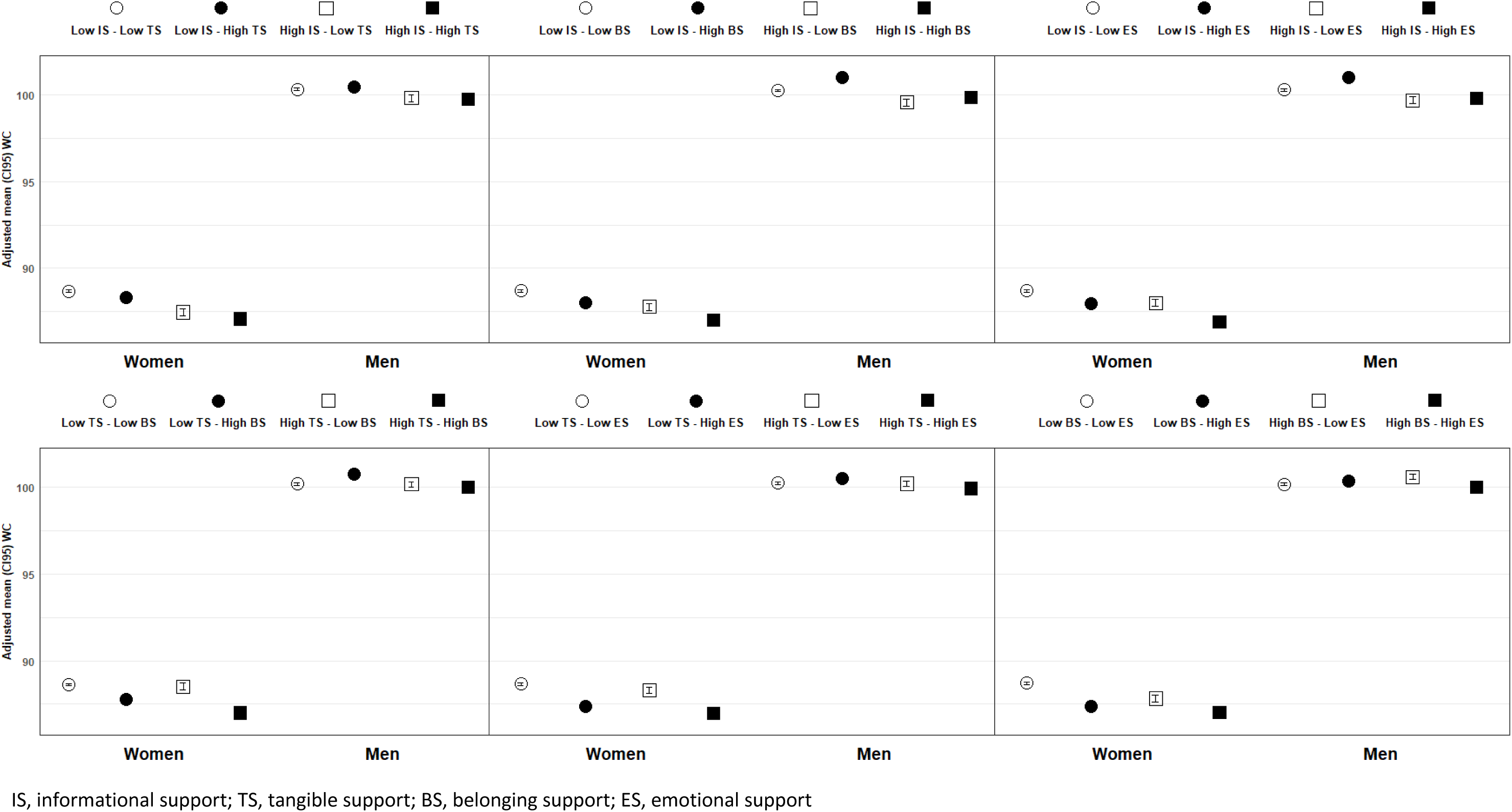
Adjusted mean WC for the interactive associations of four types of perceived social support in women and men

Among men, the largest difference in anthropometric measures was seen for the combination of informational and tangible support.

No interactive association of dual support with the odds of obesity was observed in either gender. Notably, men with low informational support had 21% higher odds of general obesity (OR 1.21 [1.06, 1.38]) and 16% higher odds of central obesity (1.16 [1.01, 1.34]) (Supplementary Table S2).

Sensitivity analyses of the final models included additional potential confounders, and results shows no appreciable changes from the main results for BMI and WC, with one exception (Supplementary Tables S5 to S8). The addition of health behaviours to our final models revealed a statistically significant interaction effect of belonging support and tangible support for WC among women as well as for general obesity (data not shown).

### Additive and interactive associations of dual social supports with blood pressure, by gender

Figure 3 shows different patterns of association between different combinations of social support and adjusted mean SBP between women and men. Among women, low availability of informational support, with or without deficits in a second support type, was associated with the highest adjusted mean SBP levels (range: 119.94 to 119.95 mmHg) (Supplementary Table S3). Only the combination of informational support and emotional support appeared to be linearly associated with adjusted mean SBP among women: that is, women with high informational and emotional support had the lowest adjusted mean SBP level (118.2 mmHg [117.96, 118.44]) than all the other high-high combinations of social support (range: 118.48 to 118.56 mmHg). Notably, higher levels of SBP were also seen among women with mixed availability of social supports, such as high belonging support combined with low emotional support (119.77 [119.41, 120.13]), high tangible support with low emotional support (119.88 [119.56, 120.19]), and high tangible support with low belonging support (120.13 [119.79, 120.48]). Among men, low-low combinations of two social supports were not consistently associated with the highest adjusted mean SBP levels, and only the combination of tangible and emotional support appeared to have a graded association with SBP. The highest mean SBP among men was observed for the combination of high belonging support and low tangible support (123.18 mmHg [122.95, 123.4]), while the lowest mean SBP among men was seen for the combination of low belonging support and high emotional support (121.40 mmHg [121.13, 121.67]).

**Figure 3.**
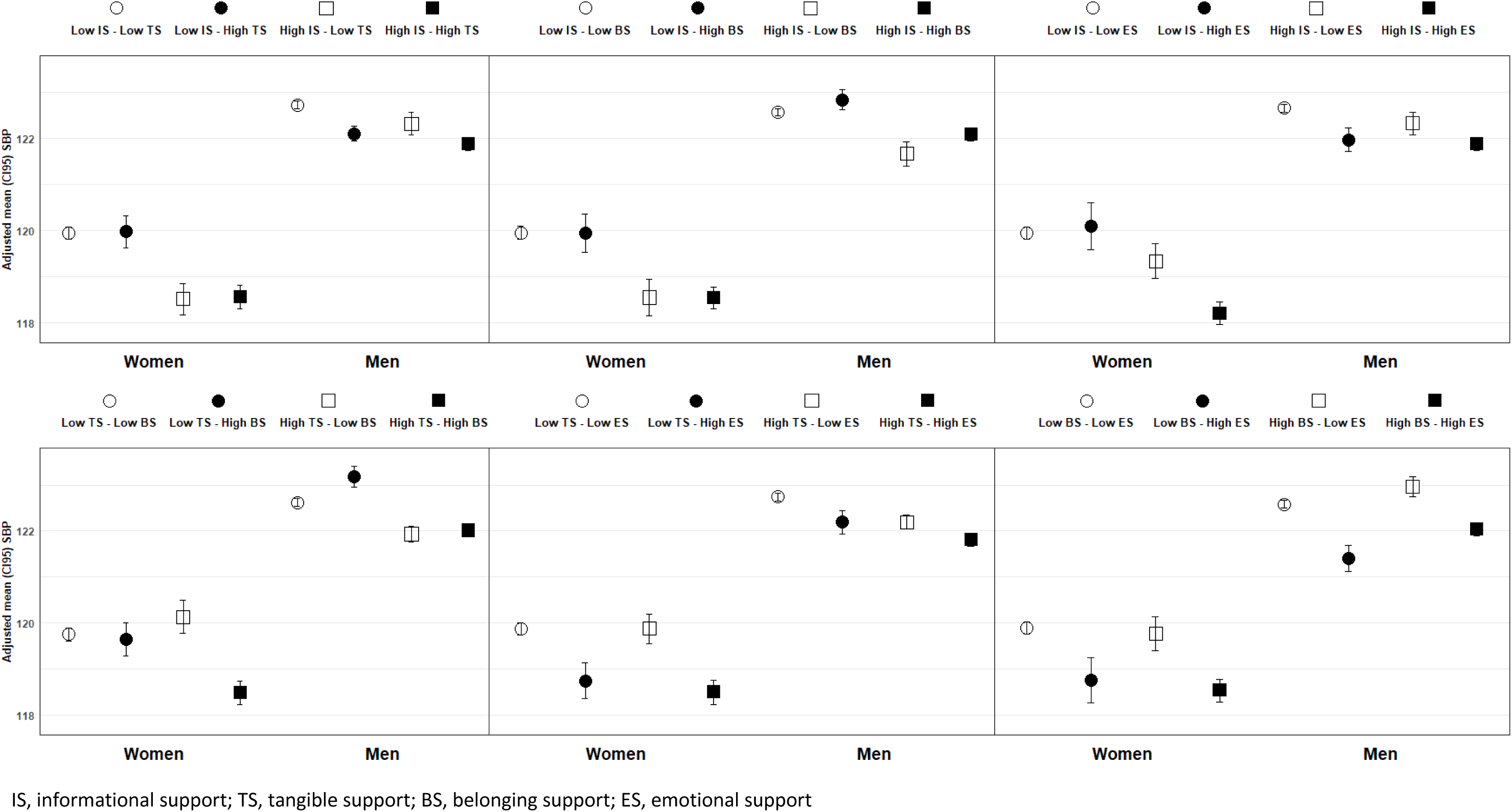
Adjusted mean SBP for the interactive associations of four types of perceived social support in women and men

Adjusted mean DBP levels appeared more similar between women and men for four of the six different combinations of social supports (Figure 4). Adjusted mean DBP levels were lowest when high tangible support combined with low informational, belonging or emotional support. The highest levels of DBP among women were observed for those reporting high emotional support combined with low tangible support (72.25 mmHg [72.15, 72.34]), low informational support (72.23 mmHg [72.1, 72.36]) and low belonging support (72.21 mmHg [72.09, 72.33]). Among men, the highest DBP levels were seen for those reporting high informational support and low tangible support (76.72 mmHg [76.5, 76.94]).

**Figure 4.**
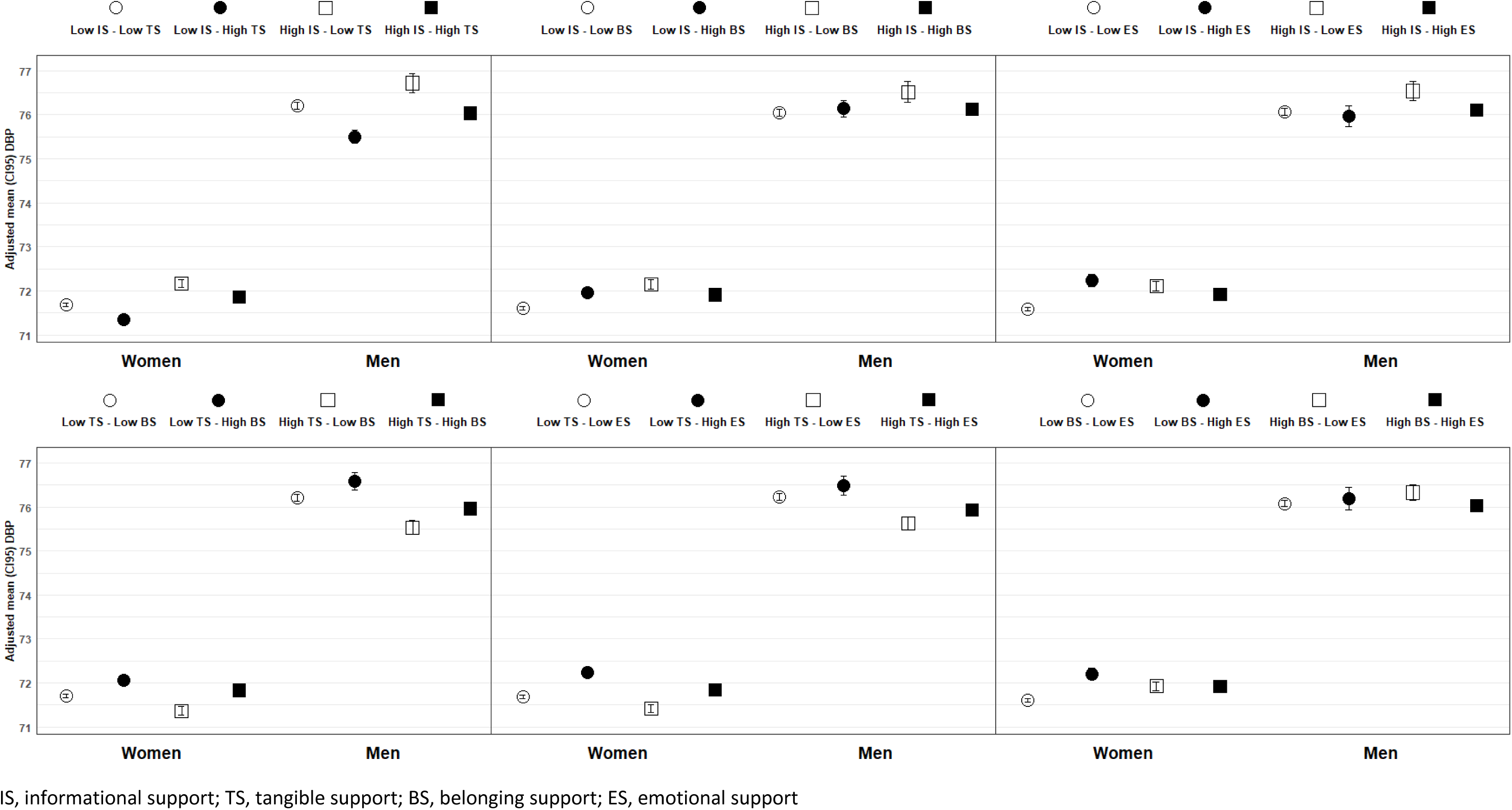
Adjusted mean DBP for the interactive associations of four types of perceived social support in women and men

No interactive association of dual support with hypertension was observed, although tangible and belonging neared significance (p<0.1) among men (Supplementary Table S4).

Sensitivity analyses of the final models included additional potential confounders, and results shows no appreciable changes from the main results reported above, with one exception (Supplementary Tables S9 to S12). The addition of psychological factors to our final models revealed a statistically significant interaction effect between belonging support and tangible support for SBP among women.

## Discussion

This population-based, cross-sectional study of a large aging cohort in Canada revealed independent additive associations between multiple social supports and CVRFs factors that differed by gender. The novel contribution of this work is the demonstration that average anthropometric and blood pressure measures were much higher for each measure of low social support when individuals also lacked a second type of social support, with consistent associations among women. Notably, our findings did not, however, support an antagonistic/synergistic effect (multiplicative interaction), meaning one form of social support did not mitigate (or exacerbate) the negative effect of another on CVRFs as some have suggested might be the case. Rather, our results point to deficits in different social supports that had additive adverse CVRF outcomes among women. Specifically, we found that all low-low combinations of social supports were associated with adjusted mean WC levels more than 88 cm that are considered ‘very high risk’ for heart disease among women. In addition, low availability of informational support, with or without deficits in a second support type, was associated with the highest adjusted mean SBP levels among women. Overall, these results indicate that women and men differed quite dramatically in the specific configurations of social support deficits that were most strongly associated with major CVRFs.

### Findings in the context of previous research

Both perceived and received social support (functional social relationships) are known to influence CVRFs in older adults,^9,25,26^ and thus social support is viewed as a critical part of comprehensive efforts to mitigate the future burden of CVDs.^25^ Notably, previous research has called for the examination of social support as a multidimensional concept relevant to CVD risk prevention.^9^ Emotional support in particular has been identified as one important dimension of social support for physiological processes of the cardiovascular, endocrine and immune systems;^9^ emotional/informational support has also been associated with incident mild cognitive impairment or dementia in older women.^27^ In addition, informational support from neighbors and emotional support from close friends were found to have an interaction with feelings of loneliness in relation to hypertension in a rural Chinese sample of middle- and older-age adults.^28^ Informational support, emotional support, belonging support and tangible support were each associated with hypertension in our sample of Canadian older adults, although associations were small and their directions were more consistent among women.^15^ Similarly, all four types of social support were associated with adiposity but only for older Canadian women and not for older men in the same cohort.^16^ Yet, the potential interplay of different types of social support as a unique determinant of CVD risk in older adults remains unexamined. This study is the first, to our knowledge, to assess inter-relations of multiple social supports with respect to two major CVD risk factors in older adults.

Limited research suggests that there is an interaction of social support with other psychosocial (e.g. optimism, social strain, stress) and/or socioeconomic resources (e.g. education, income) in relation to cardiovascular-related health outcomes. A study of a US birth cohort showed that social support was a positive psychosocial resource that predicted a healthy adult body mass index and buffered the effects of childhood socioeconomic disadvantage.^29^ Among adult partners, supportive marital relationships buffered the negative association between income and ambulatory diastolic blood pressure.^30^ The joint effects of social support and education on blood pressure among African American adults was also shown: greater received social support was positively associated with higher systolic blood pressure among individuals with low education levels.^31^ In a small community sample of older adults, emotional social support was a coping resource that both predicted depressive symptoms and also moderated the impact of stress on depressive symptoms.^10^ Main effects, stress-buffering and joint effects were also observed for social support and psychological wellbeing in Mexican-origin adults, with high social support buffering against the negative association of life stress on psychological wellbeing.^12^ However, while most research indicates that social support and other social-level resources have a joint effect on cardiovascular health, some findings show that the moderating role of social support in hypertension may not buffer social stressors in all settings.^28^ The synergy effects reported for health outcomes do not mirror this study’s findings of an additive effect that the average anthropometric and blood pressure measures were much higher for each measure of low social support when women also lacked a second type of social support. The only significant interactive effects were observed in sensitivity analyses of WC and SBP among women, with specific covariables added (health behaviours and psychological factors, respectively).

Our results indicated that women and men were differentially vulnerable to greater CVD risk from distinct combinations of poor social supports. Overall, findings demonstrated that women fared worse in their CVD risk profiles in the absence of both informational support and a second type of social support. This finding was more notable for BMI and WC than for BP. By contrast, among men, there was only one clear association between two absent social supports and CVRFs. The direct effect of social support and its interactions with other social factors on BP has been shown to differ for women than men. In a small community study of about 200 adults aged 21 to 50 years, social support was examined in relation to blunted nocturnal dipping as an independent predictor of cardiovascular morbidity and mortality; results showed that normotensive adults with low levels of social support had blunted dipping with greater benefits of social support seen among women than among men.^32^ Another study of 2348 married/co-habiting adults aged 25 to 75 years showed that supportive networks could buffer the insalubrious effects of strained interactions on wellbeing and health outcomes, but the buffering role of friends and family was observed more frequently for women than for men.^13^ Although the direct effects of social support on cardiovascular-related outcomes appears stronger or more consistent among women adults across the age spectrum, the effects of social support on cardiovascular-related behaviours may be stronger for men. Recent research from Spain showed that middle-and older-age men who lack social support have the lowest adherence to cardiovascular screening (e.g. self-reported testing of blood pressure) and lifestyle recommendations but older women have high adherence irrespective of social support.^33^ Thus, it is clear that sex-and-gender-based differences exist in the relationship between social support and CVD risk and this study indicates that the additive interplay of social support with another social factor is more strongly associated with objectively measured anthropometry and blood pressure in older women.

### Strengths and limitations

Although this study is cross-sectional and thus prohibits any causal inference, it provides information on which associations to examine prospectively. Self-reported measures of perceived social support can be subject to recall or survey effect bias, however multiple outcomes were clinically assessed. As an observational cohort study, results may be biased due to unmeasured confounding from measures not included; in particular, this study did not include lipids as a key CVRF as blood lipid data were not available for research in the CLSA dataset. There is also likely to be residual confounding from imprecise measurement of self-reported covariables such as health behaviours (‘lifestyle factors’) that could introduce bias that would either attenuate or inflate observed associations;^34^ these factors could also be potential mechanisms underlying the association of social support and CVRFs. Finally, our study’s external validity is limited by the nature of the CLSA cohort that is predominantly cisgender, heterosexual, White adults. Nevertheless, our focus on (cisgender) women provided important disaggregated data that could reveal how the role of social support for CVRFs may be gendered from the lens of power relations between women and men.^35^ Finally, our results can only be generalized to middle- and older-age adults living in the community in similar high-income countries.

There are multiple notable strengths of this population-based study which include: a large sample size, adjustment for multiple known confounders, sex-and-gender-based analysis (SGBA), a focus on interaction effects, and several objectively measured cardiovascular risk factors. This study explicitly focused on the joint associations of four types of functional aspects of social relationships rather than using a summary index that masks the relative contribution of each type of connection. A particular strength of our study is in its consideration of the gender differences in this more complete picture of different social supports and potential synergy that matter for cardiovascular health. Attention to both gender and the multiple mutually reinforcing social supports thereby adds more precision to the cardiovascular prevention literature that has implications for both research and clinical practice.

## Conclusions

This study demonstrated that the combination of two deficits in social support showed much worse CVD risk profiles than a single deficit for women only. Only informational support alone was associated with obesity among men. Overall, results contribute empirical knowledge on the additive effect and lack of an antagonistic interactive effect between multiple social supports for CVRFs. Care and prevention of CVD in older women would benefit from addressing several specific types of social support, including informational support.

## Data Availability

Due to privacy and confidentiality requirements data can only be made available for researchers who meet the criteria for access. See Canadian Longitudinal Study on Aging (CLSA) https://www.clsa-elcv.ca/data-access for details

## Acknowledgement

This research was made possible using the data/biospecimens collected by the Canadian Longitudinal Study on Aging (CLSA). Funding for the Canadian Longitudinal Study on Aging (CLSA) is provided by the Government of Canada through the Canadian Institutes of Health Research (CIHR) under grant reference: LSA 94473 and the Canada Foundation for Innovation, as well as the following provinces, Newfoundland, Nova Scotia, Quebec, Ontario, Manitoba, Alberta and British Columbia. This research has been conducted using the CLSA Baseline Comprehensive Dataset version 4.0, under Application Number 19CA003. The CLSA is led by Drs. Parminder Raina, Christina Wolfson and Susan Kirkland.

## Funding disclosure

This secondary data analysis study was funded by the Canadian Institutes of Health Research (grant #162987). The funders had no role in study design, data collection and analysis, decision to publish, or preparation of the manuscript.

## Disclaimer

The opinions expressed in this manuscript are the author’s own and do not reflect the views of the Canadian Longitudinal Study on Aging.

## Sources of support/funding

The Canadian Institutes of Health Research Catalyst Grant; Faculty of Pharmaceutical Sciences start-up funds for hardware and software.

## Conflicts of interest

none declared

## Notes

### Competing Interest Statement

The authors have declared no competing interest.

### Funding Statement

Yes

### Author Declarations

UBC Behavioural Research Ethics Board (H19-00971).

